# The temporal correlation between positive testing and death in Italy: from the first phase to the later evolution of the COVID-19 pandemic

**DOI:** 10.1101/2021.07.14.21260505

**Authors:** Vincenzo d’Alessandro, Nicole Balasco, Pietro Ferrara, Luigi Vitagliano

## Abstract

After the global spread of the novel coronavirus disease 2019 (COVID-19), research has concentrated its efforts on several aspects of the epidemiological burden of pandemic. In this frame, the presented study follows a previous analysis of the temporal link between cases and deaths during the first epidemic wave (Phase 1) in Italy (March-June 2020). We here analyze the COVID-19 epidemic in the time span from March 2020 to June 2021. The elaboration of the curves of cases and deaths allows identifying the temporal shift between the positive testing and the fatal event, which corresponds to one week from W_2_ to W_33_, two weeks from W_34_ to W_41_, and three weeks from W_42_ to W_67_. Based on this finding, we calculate the Weekly Lethality Rate (WLR). The WLR was grossly overestimated (~13.5%) in Phase 1, while a mean value of 2.6% was observed in most of Phase 2 (starting from October 2020), with a drop to 1.4% in the last investigated weeks. Overall, these findings offer an interesting insight into the magnitude and time evolution of the lethality burden attributable to COVID-19 during the entire pandemic period in Italy. In particular, the analysis highlighted the impact of the effectiveness of public health and social measures, of changes in disease management, and of preventive strategies over time.

## Introduction

The novel Coronavirus Disease 2019 (COVID-19) pandemic that started in China in the last months of 2019 is causing a global health emergency, the solution of which seems yet far to be achieved despite the enormous efforts made to mitigate its spread and develop therapeutic and effective preventive strategies (1–3). Currently, the official number of cases (i.e., infected subjects) is approaching 200 million with nearly four million deaths (4,5). The pandemic has non-uniformly affected almost all continents and countries (4), often with a fast and intricate evolution resulting from the combination of many factors, namely, (a) the use of personal protective equipment, (b) restrictions to the people mobility applied by public authorities that were periodically released in a difficult attempt to prevent or limit the infections without destroying local economies, (c) the seasonality of viral respiratory diseases, (d) the progressive appearance of SARS-CoV-2 variants endowed with increased infection rates, and (e) the recent implementation of vaccination campaigns (2,3,6,7).

In this scenario, the gathering of information on the burden of the disease is of utmost importance for better understanding the origin and evolution of the pandemic spreading and the effectiveness of the public health interventions (3). By analyzing the evolution of the pandemic outbreak in Italy, the center of the first main outbreak among Western countries, we have recently examined the temporal correlation between the positive testing to COVID-19 and deaths in the first phase, i.e., the period March-June 2020 (8,9). In particular, we found, on average, a one-week delay between the positive test and the fatal event. Despite the straightforwardness of the approach and the heterogeneity of the available data, we exploited this finding to propose a lethality measure denoted as Weekly Lethality Rate (WLR), which is based on the ratio between the number of deaths occurring in a certain week and the number of positive tests detected in the previous weeks (8).

In order to further investigate the lethality impact of the infection in Italy, we have extended this analysis to the entire pandemic, by monitoring the temporal link between positive testing and death from the summer 2020 to June 2021. As for the initial phase, we quantified the temporal correlation between these events, and found a progressively increasing time gap. On this basis, we conceived an approach to generalize the WLR. The implications of these findings on the evolution of the Italian outbreak will be discussed, considering the impact and effectiveness of political interventions, changes in disease management, and preventive strategies.

## Results

### Comparative analysis of the evolution of cases and deaths

The inspection of the daily deaths/cases curves (**Figures 1** and **S1**) clearly indicates that the country suffered from two main outbreaks (here denoted as Phase 1 and Phase 2). The first started at the end of February 2020 and ended in the summer of the same year. Moreover, starting from the fall 2020 a second remarkable increase of both cases and deaths was experienced. Although this second phase is progressively regressing, it is still ongoing (June 2021). Notably, the maxima of the deaths curve are pretty similar in the two phases while the maximum of the cases is significantly higher in the second phase compared to the first one.

**Figure 1.**
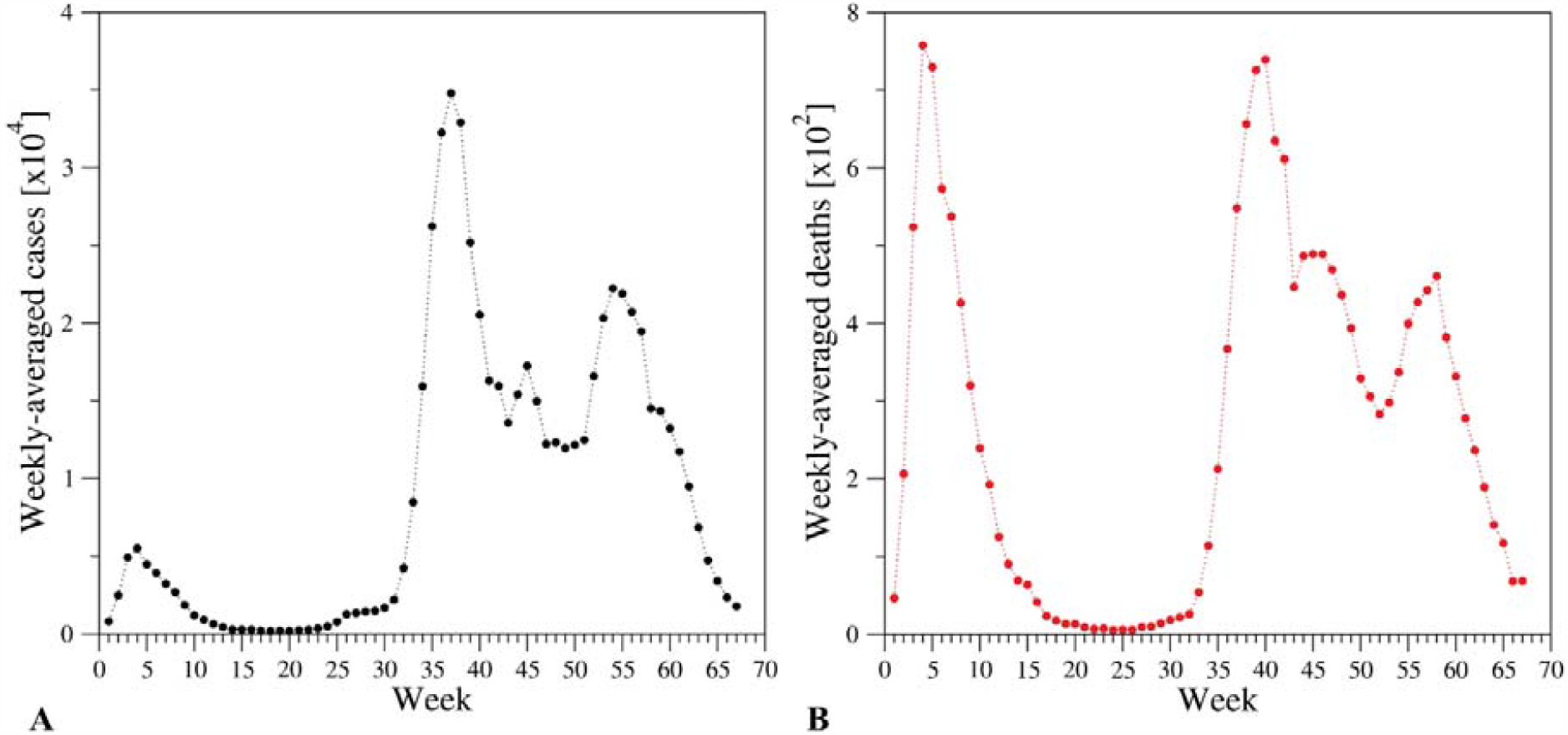
Evolution of the weekly-averaged (A) cases (positive tests) and (B) deaths. Weeks have been numbered according to Table S2.

As previously explained, the application of the temporal 1-week shift scheme that was successfully exploited to analyze Phase 1 (8) (**Figure 2A**) produced a very poor fitting of the curves of Phase 2 (**Figure 2B**). Better fittings are obtained by using shifts of two or three weeks to the curve of the cases (**Figure 2C and 2D**). A closer inspection of the fitting clearly indicates that a unique shift scheme cannot account for the complexity of Phase 2. This observation is not surprising considering the remarkable time span of Phase 2 (~9 months). A similar conclusion is reached when the sum of squared residuals (SSR) between the normalized daily cases/deaths is calculated upon the systematic shifts of the curve of the cases of Phase 2. As shown in **Figure S2**, the SSR values present a marked weekly periodicity due to the daily dependence of testing and death registrations. Interestingly, two nearly identical global minima corresponding to shifts of either 13 or 20 days are evident. In this scenario, considering the complexity of Phase 2, we deconvolute the global cases/deaths curves by identifying the underlying curves corresponding to the three subphases (2.1, 2.2., and 2.3) (**Figure 3A** and **Figure 4A**). This was done by noticing that the ascending parts of the curves can be described by sigmoidal increases whereas the descending regions are characterized by exponential decreases (see Methods for details). In this framework, the main parameter for the exponential function was derived from the fitting of deaths of the descendent region of Phase 1.

**Figure 2.**
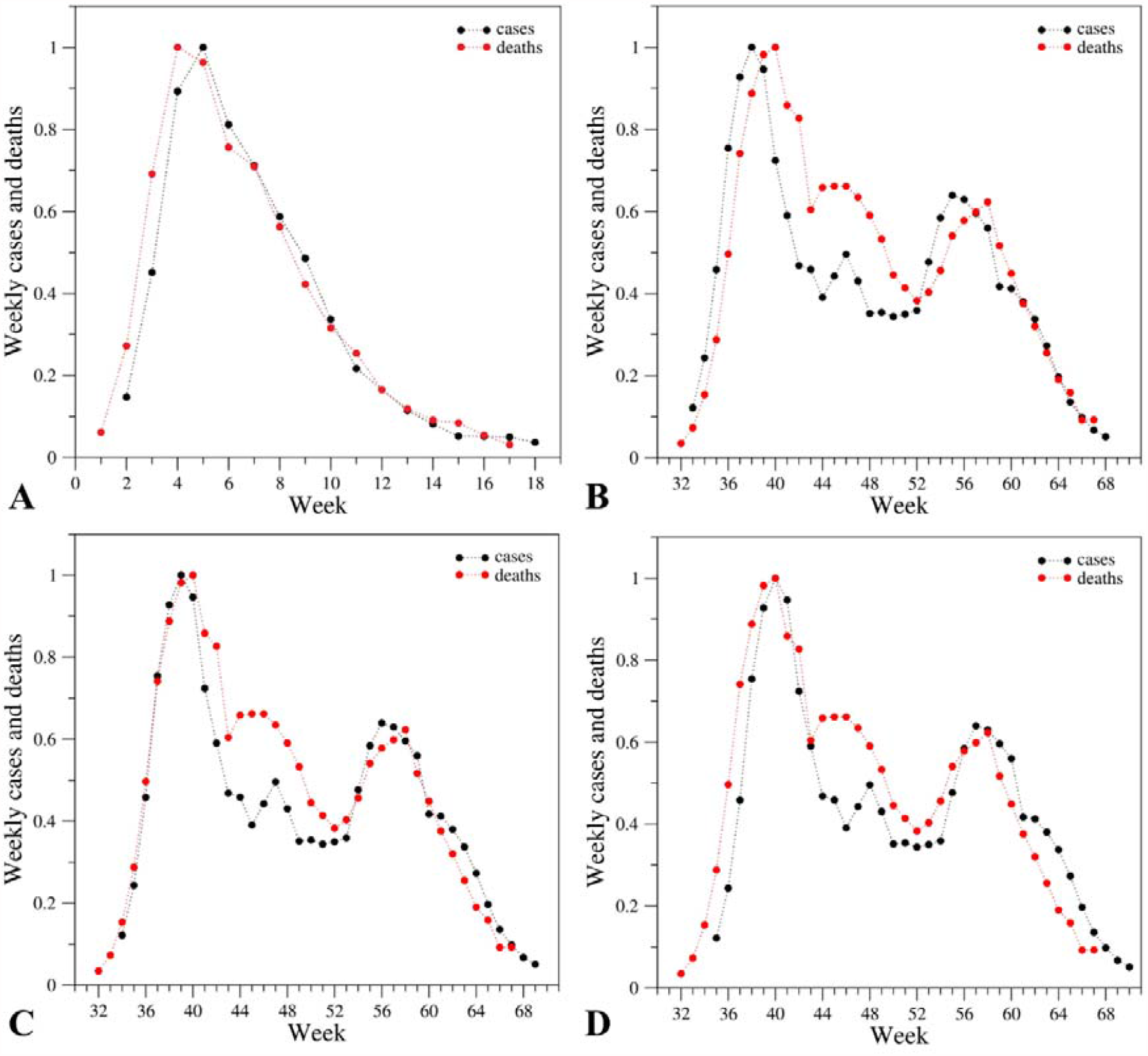
Comparison of the evolution of the weekly cases (black) and deaths (red) upon normalization of the curves in (A) Phase 1 (W_1_-W_17_) and (B-D) Phase 2 (W_32_-W_67_) of the pandemic. The normalization was performed by dividing the actual values by the maximum of each ensemble. The curve of cases is one-week (A, B), two-week (C), three-week (D) shifted ahead.

**Figure 3.**
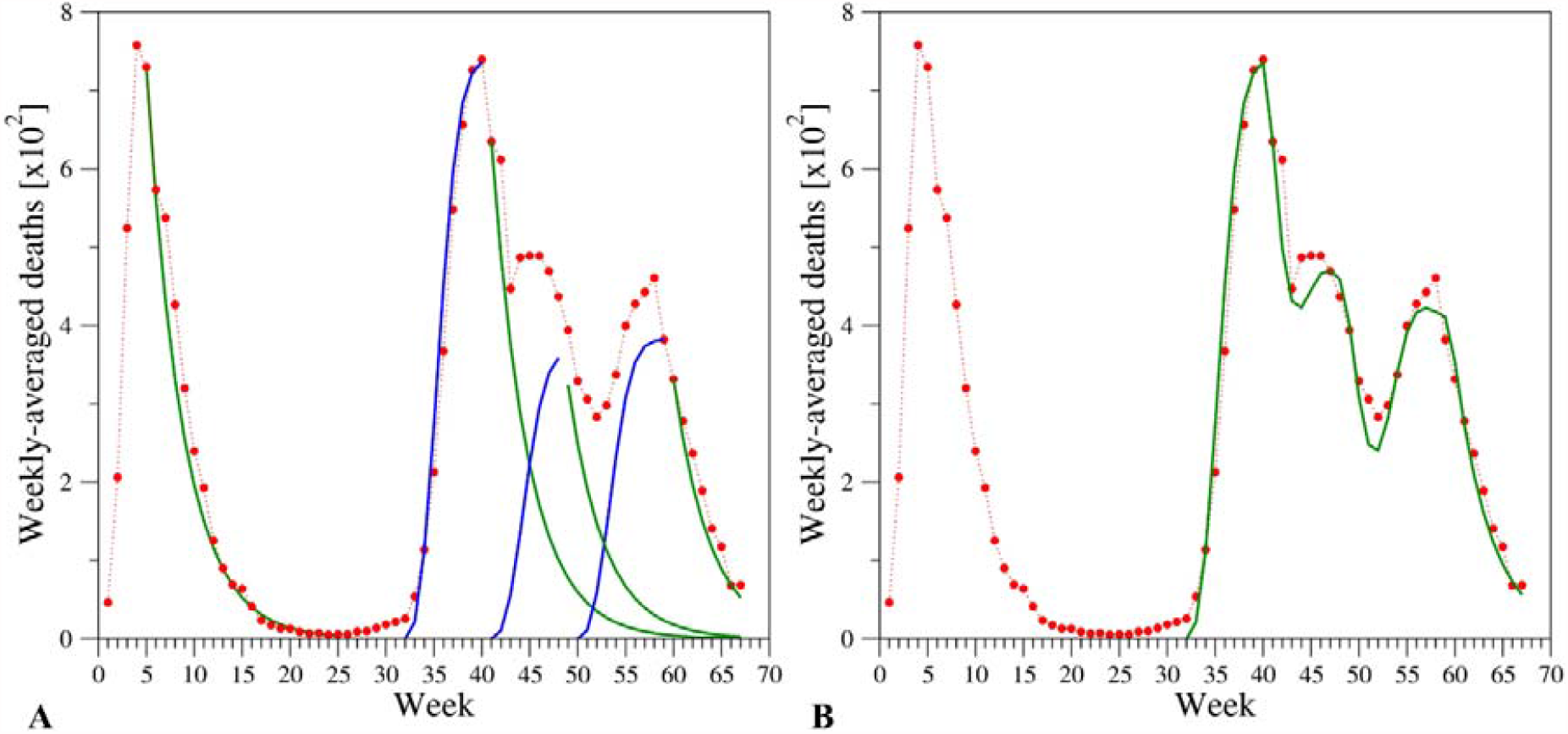
(A) Modeling of the three subphases of the curve of deaths of Phase 2 with mathematical functions. For the rises, sigmoidal functions making use of exponent 2.5 and time constant 4.1 were exploited. The time constant 3.8 calibrated for the fall of deaths occurring in Phase 1 was also employed for the falls of the three subphases of Phase 2. (B) Model (sum of the 3 modeled subphases) of the second wave superimposed to the real data.

**Figure 4.**
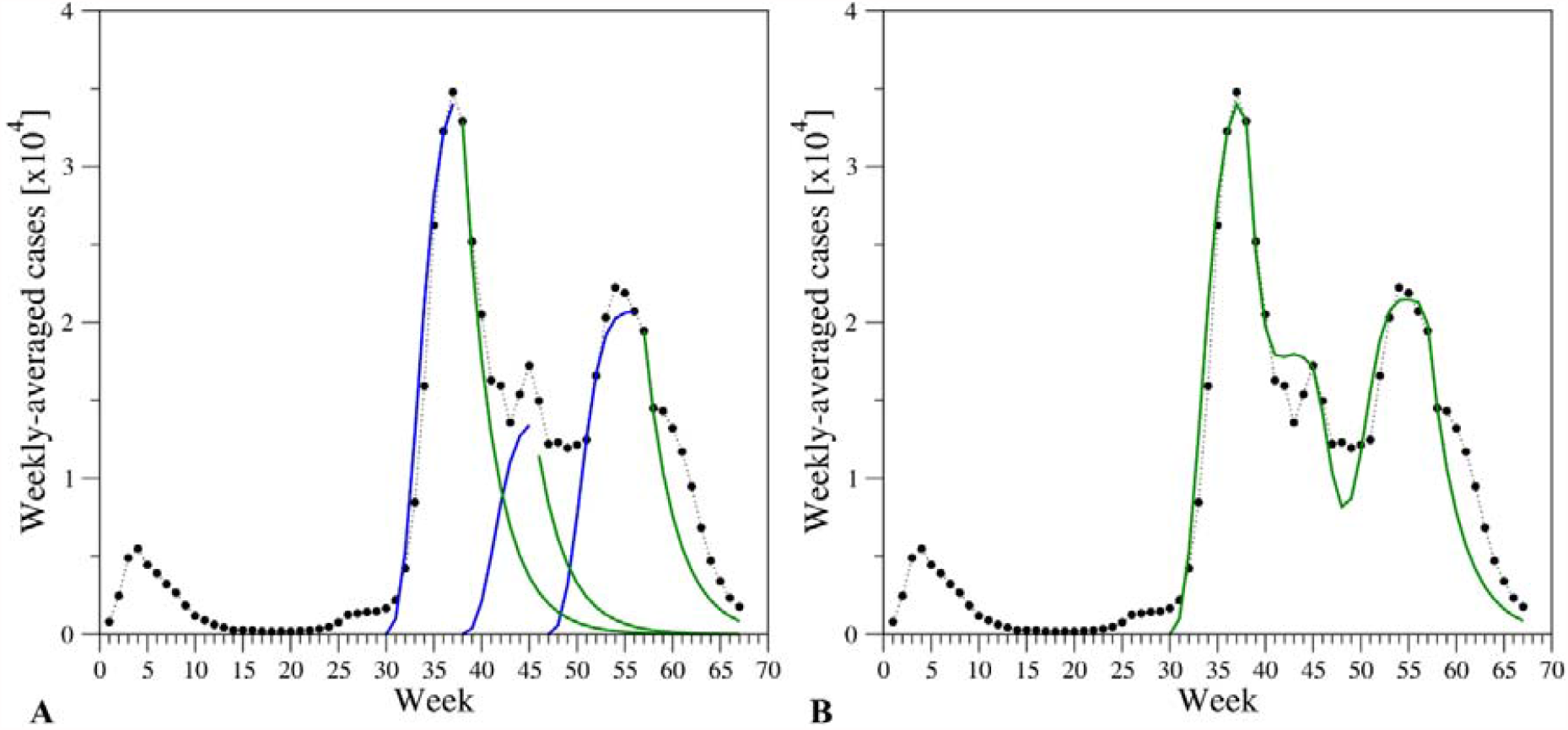
(A) Modeling of the three subphases of Phase 2 of the curve of cases with mathematical functions. For the rises, sigmoidal functions making use of exponent 2.5 and time constant 4.1 were exploited. The time constant 3.2 was employed for the falls of the three Phase 2 subphases. (B) Model (sum of the 3 modeled subphases) superimposed to the real data.

The superposition of the deconvoluted curves (**Figure 3B** and **Figure 4B**) shows a remarkable agreement with the real cases/deaths. The only significant discrepancies are observed in the time interval W_42_-W_45_ that corresponds to mid-December – mid-January when, due to the holiday season, the recording of cases and deaths was occasionally postponed.Such a deconvolution also provides a measure of the temporal shift between the positive testing and the fatal event. An inspection of the equations used to fit the experimental data indicates that the rise of the deaths in Subphase 2.1 is shifted by two weeks from the corresponding positive testing; in the fitting curve, the parameter W_32_ or W_30_ is indeed present for deaths and cases, respectively. On the other hand, for the subsequent weeks the equations suggest a 3-week shift since the parameter in the equations is either W_41_ (deaths) or W_38_ (cases). These observations are confirmed by the indications provided by the SSR analysis described above.

### Weekly Lethality Rate

Based on the variable shifts between the curves of deaths/cases detected throughout Phase 2, we adapted the definition of the WLR previously introduced (8) by alternatively considering a single week (from W_2_ to W_33_) shift, a 2-week shift (from W_34_ to W_41_), or a 3-week (from W_42_ to W_67_) shift. As shown in **Figure 5** and **Table S4**, we observed significant changes of this parameter in the different stages of the pandemic. In particular, the value of WLR was grossly overestimated (~13.5%) in Phase 1 (8). This is likely due to the severe underestimation of the cases at that time. A WLR value of about 2.6% was observed in most of Phase 2. Interestingly, a slow but significant reduction of this parameter has taken place in the last weeks (W_50_-W_67_; February 8^th^ – June 13^th^, 2021). In particular, at W_67_ the WLR assumes a value (1.4%) that is almost halved when compared to the nearly constant one detected in the period W_34_-W_49_.

**Figure 5.**
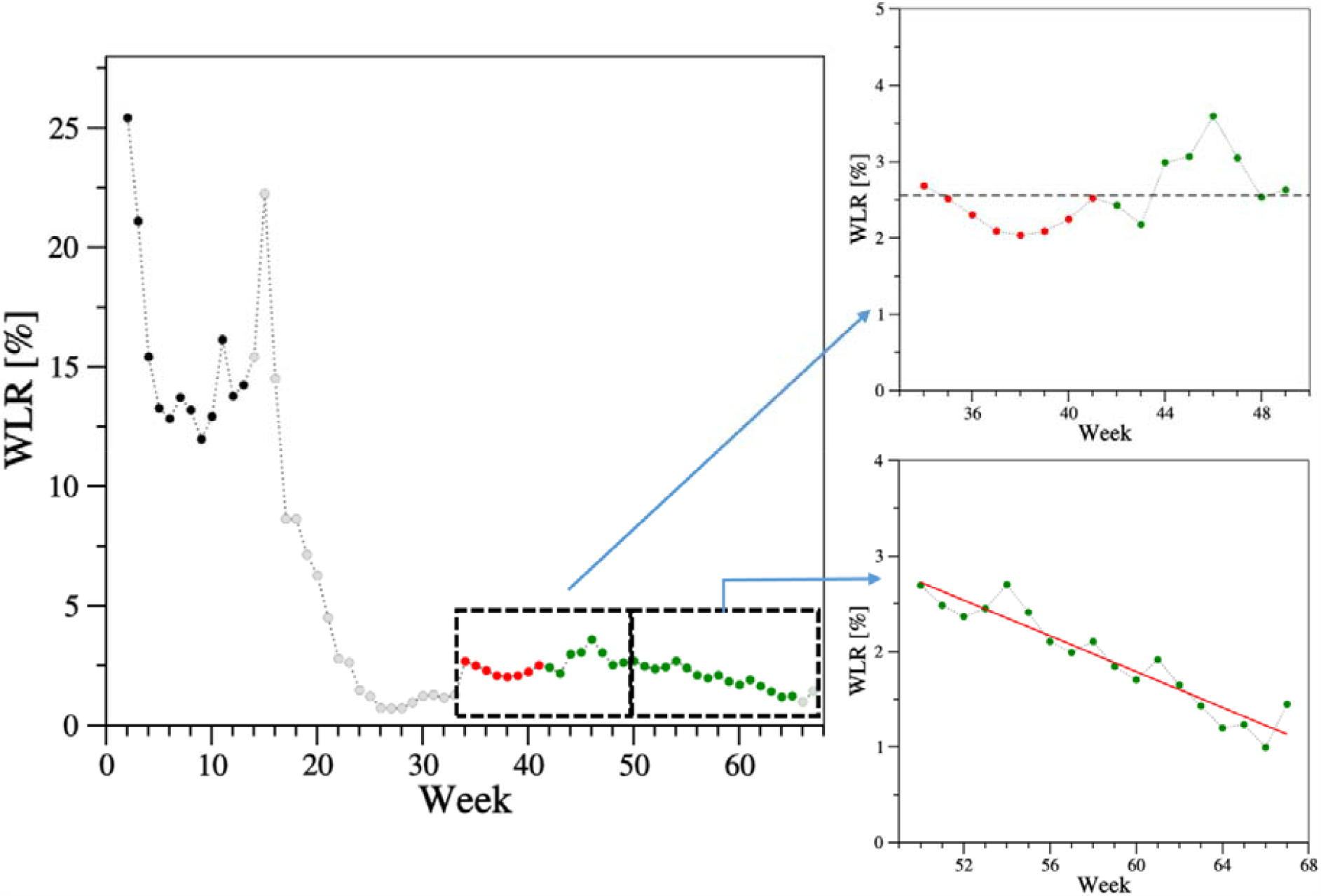
Weekly lethality rate (WLR) evolution in the overall pandemic. The WLR values were calculated applying a 1-week (black and grey), 2-week (red), and 3-week (green) shift between positive test and death in the time intervals W_2_-W_33_, W_34_-W_41_, and W_42_-W_67_, respectively. Values computed for the weeks (W_14_- W_33_, W_66_-W_67_) with less than 70 weekly-averaged deaths are in light grey. The average WLR value (2.56%) detected in the time interval W_34_-W_49_ is shown as a dashed line. The linear regression analysis of the WLR values in the weeks W_50_-W_67_ provides a correlation coefficient R=-0.95 (p-value<10^−5^). The regression line (y = 7.4091 - 0.093675 x) is shown in red.

## Discussion

One of the most striking and worrying features of the COVID-19 pandemic is its unpredictable evolution: with the exception of an extremely limited number of nations that have been able to control the infections, the vast majority of the countries have suffered from phases characterized by a high COVID-19 burden that was alternated with time periods in which the pandemic was essentially under control. This articulated evolution has also been experienced in Italy, the center of the first main outbreak across Western countries. In the initial stage of the pandemic (February – June 2020), quite strong measures were taken by the governmental authorities to severely limit the mobility of people and therefore the diffusion of the infection (2,8). Despite the gross underestimation of cases in the early months of the outbreak (due to the emergency phase and a low capacity of case detecting), the enforcement of severe lockdown restrictions contributed to the clear-cut shape of the curves, with an ascending curve followed by a monotonic descending one (**Figure 1**). The analogous trends displayed by the curves reporting cases and deaths prompted us to search for a temporal link between them. A remarkable good fitting between these two profiles was obtained when the curve of the cases was shifted by one week (8). The well-defined quantification of this temporal link provided us the opportunity to define a time-dependent WLR that was nearly constant in the first months of the pandemic. The inspection of the WLR profile indicates that, after assuming rather large values (~13.5%) in Phase 1 due to the underestimation of cases, it decreased to very low values (below 1%) during the summer of 2020 (**Figure 5**). Although the marked reduction in the number of cases/deaths observed in this period makes the calculated WLR less reliable, its drop may be ascribed to different factors including the lockdown measures, the seasonality of viral respiratory diseases, and an increased virus circulation amongst young people who are less susceptible to severe and fatal COVID-19 outcomes (10,11). Therefore, after the initial outbreak and the summer months, when the infections were essentially under control, from autumn Italy suffered a second phase (Phase 2) of the pandemic that is still ongoing. Differently from the first, Phase 2 has been characterized by an intricate evolution with multiple rises and decreases of the infections. On the basis of the peaks that we observed in the curves of both cases and deaths, we were able to identify and model three distinct subphases. As a whole, the shape of the Phase 2 curve mirrored the differences in virus circulation and type of restrictive public health measures compared to Phase 1. From an epidemiological standpoint, in the first half of 2020, the major spread of SARS-CoV-2 was limited to northern regions of the country, which registered the biggest COVID-19 outbreak in terms of both cases and death toll (12); from September 2020, the virus has been circulating across the entire country (10). Again, in order to reduce the social and economic consequences of protracted lockdown restrictions, governmental authorities adopted less-stringent limiting measures, the impact of which was reflected in the curves. A system of region-based risk levels was implemented, which allowed the easing of measures according to a set of indicators (for instance, number of cases and deaths, number of intensive care unit [ICU] admissions, percentage of occupied ICU beds, etc.), with different extents of SARS-CoV-2 circulation across regions and weeks (10). Interestingly, despite the heterogeneity of the data that are separately collected in the twenty-one regions/territories of the country and the complexity of Phase 2, we could quantify the temporal link between cases and deaths. Notably, the shift between the positive testing and the fatal event is longer than that observed in Phase 1 (one week) and is progressively increasing during Phase 2 (two or three weeks). This difference can be ascribed to the improved timeliness of the testing, which was only reserved to people displaying multiple symptoms in Phase 1 and, possibly, to some improvement in the therapeutic interventions that delayed the death in Phase 2. Moreover, the identification of this temporal link gave us the opportunity to evaluate the lethality attributable to COVID-19 in Italy during the whole epidemic period, from March 2020 to June 2021, using complete epidemiological data of SARS-CoV-2 spread in the country. A significant increase of the WLR took place in October 2020, concurrently with a new increase of the virus spread and its following circulation in susceptible populations, like elder individuals or people in fragile states (14). From the beginning of Phase 2 to the beginning of February 2021 the WLR assumed a rather constant value (~2.6%) (**Figure 5**). Starting from mid-February 2021 (W_51_), the WLR is significantly decreasing. It is important to note in this time interval the WLR practically halved from 2.6 to 1.3%, the value detected at the beginning of June 2021. Notably, this period also coincides with the progressive increase of the vaccination coverage in Italy, primarily in at-risk and susceptible subjects, with a consequent positive impact on COVID-19 burden and lethality reduction (7). Further research is needed to better investigate the impact of vaccines and vaccination campaign on SARS-CoV-2 diffusion and lethality.

Some limitations of our study should be acknowledged. First, as for our previous analysis of lethality during the first epidemic wave (8), the research included information measured through surveillance systems where data were provided in aggregated form and without any case stratification; thus, it was not possible to evaluate uncertainty sources and adjust results for potential independent predictors of death, but this research was intended as a straightforward strategy to assess the time-trend of the proportion of cases who died from the disease. Second, the WLR uses the number of subjects that tested positive as population (denominator), thus being influenced by the number of tests performed on a certain time-point. However, the combination of data on a weekly-aggregated manner greatly reduced possible differences across different week-days. Lastly, even if the 1- to 2-week and 2- to 3-week shifts were based on static denominators, the WLR calculated a ratio representing relative rates of deaths which reflect analytical expressions of lethality proxy over time.

In conclusion, our study provides interesting insights into the evolution of the COVID-19 pandemic in Italy by highlighting some distinctive features, in terms of trend complexity of the lethality rate in the different phases of the pandemic. Our approach also documented the impact of the public health measures on SARS-CoV-2 spread and associated lethality, also highlighting the possible positive effect of vaccination efforts, but more studies assessing this hypothesis should be implemented in the near future. The application of the proposed approach could help examine data from other contexts, allowing comparison with the lethality associated with SARS-CoV-2 in other countries.

## Materials and Methods

### Study Design and Data Source

We performed a longitudinal retrospective analysis on the time-trend of the lethality due to COVID-19 in Italy through the data from the freely accessible national COVID-19 integrated surveillance system (13). More specifically, we gathered the daily number of diagnostic tests, confirmed cases, and deceased related to SARS-CoV-2 (**Table S1**). We traced data over 67 weeks (denoted as W_1_, W_2_, …, W_67_) covering the period from March 2^nd^, 2020 to June 13^th^, 2021 (**Table S2**).

### Statistical Analysis

Numbers of cases, deaths, and tests were grouped in a week-based manner (**Table S3**). The average daily values (also denoted as weekly-averaged values) of cases and deaths were obtained by dividing the total weekly number by seven.

Following our previous study of the time evolution of COVID-19 lethality during the first wave (8,9), our analysis included:

(i) Identification of the different phases of the epidemic. On the basis of the inspection of the cases and deaths curves, we conventionally set the end of the first phase on June 28^th^, 2020, latest day of week 17 (W_17_); hence, the first phase (also referred to as Phase 1) lasted 17 weeks (119 days). Similarly, the beginning of the second phase (Phase 2) was set on October 5^th^, 2020, first day of W_32_; thus, the second phase has lasted 36 weeks so far (252 days).

(ii) Dissection of the data and analytical modeling of the detected cases/deaths: we first described the lockdown-driven fall of Phase 1 and the initial rise of Phase 2 with a decreasing exponential and a sigmoidal function, respectively. Exponential and sigmoidal functions with calibrated parameters were then used for the falls and rises of all other curves.

(iii) The temporal shift of the cases/deaths curves was obtained by examining the derived functions. In addition, we performed a sensitivity analysis relying on the sum of squared residuals (SSR) in order to evaluate the quality of the fitting between the aforementioned curves upon specific shifts.

(iv) Depending on the detected shift at different phases/subphases of the pandemic, the WLR values were calculated by dividing the average daily number of deaths of a given week (W_i_) by the average daily number of cases of one week (W_i-1_), two weeks (W_i-2_), or three weeks (W_i-3_) before. In detail, the WLR values were computed applying a 1-week, 2-week, and 3-week shift between positive test and death in the time intervals W_1_-W_33_, W_34_-W_41_, and W_42_-W_67_, respectively.

In order to detect the temporal link between positive testing to the virus and death (as done for Phase 1 (8)), we partitioned Phase 2 into three distinct subphases, referred to as 2.1, 2.2, and 2.3, defined on the basis of the peaks detected in the cases/deaths curves on Nov 13^th^/Dec 3^rd^, 2020, Jan 6^th^/8^th^, 2021, and Mar 12^th^/Apr 9^th^, 2021 (**Figures 1** and **S1**), respectively.

In our *post-hoc* sensitivity analysis, the application of the temporal 1-week shift scheme – successfully applied to analyze Phase 1 (8) (**Figure 2A**) – produced a very poor fitting between the cases and deaths curves of Phase 2 (**Figure 2B**). Better fittings are obtained by applying shifts of two or three weeks to the curve of the cases (**Figure 2C and 2D**). A closer inspection of the fitting clearly indicates that a unique week shift scheme cannot account for the complexity of Phase 2.

The analytical modeling of the weekly-averaged cases/deaths in Phase 2 was performed as follows. As a first step, the lockdown-driven fall of the deaths of Phase 1 was favorably described with a decreasing exponential (**Figure S3**)

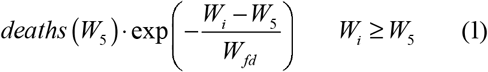

the time constant W_fd_ of which was calibrated to 3.8 weeks to obtain the best agreement between (1) and the real data.

The initial deaths rise of Subphase 2.1 was described with the sigmoidal function (**Figure S3**)

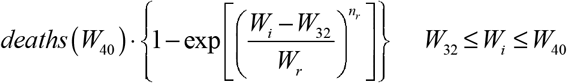

where the time constant W_r_ and power factor n_r_ were adjusted to 4.1 weeks and 2.5, respectively (**Figure S3**); deaths(W_40_) represents the peak value of this subphase.

Subsequently, exponential and sigmoidal functions with the same values for W_fd_, W_r_, and n_r_ were adopted also to model the deaths falls of Subphases 2.1, 2.2, 2.3, as well as the rises of Subphases 2.2 and 2.3, respectively (**Figure 3A**). As an example, the fall of Subphase 2.1 was described with

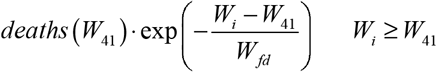

and the rise of Subphase 2.2 with

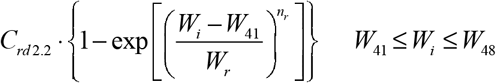

C_rc2.2_ being a fitting parameter tuned to ensure the best matching between the overall model and real data (**Figure 3B**).

A similar strategy was used to model the evolution of the weekly-averaged cases. First, it was noted that the rise of Subphase 2.1 was accurately described by the same sigmoidal function (and same parameters) exploited for the deaths (**Figure 4A**), i.e.,

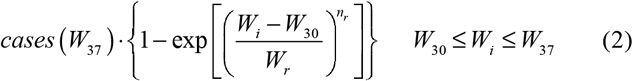

cases (W_37_) being the peak reached during this subphase. Equation (2) was also used to model the rises of cases of Subphases 2.2 and 2.3 (**Figure 4A**); as an example,

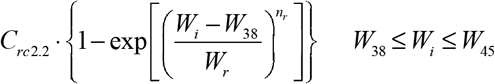

was employed for Subphase 2.2, C_rc2.2_ being a fitting parameter.

A reasonably faster time constant W_fc_=3.2 weeks was adopted in the decreasing exponentials used to describe the falls of cases in Subphases 2.1, 2.2, and 2.3 (**Figure 4A**); as far as Subphase 2.1 is concerned, the exponential function is

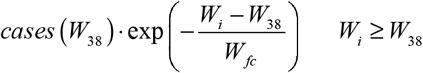

The deconvolution of the deaths/cases curves also provides a measure of the temporal shift between the positive testing and the fatal event. A comparison of the equations used to fit the experimental data indicates that the rise of the deaths in Subphase 2.1 is shifted two weeks ahead with respect to the corresponding testing: weeks W_32_ and W_30_ are indeed used to identify the rise onsets of deaths and cases, respectively. On the other hand, for the subsequent subphases the equations suggest a 3-week shift; as far as subphase 2.2 is concerned, weeks W_38_ and W_41_ are adopted for cases and deaths. These results are in line with those obtained through a comparative analysis performed by shifting the normalized curve of daily cases with respect to the deaths counterpart and calculating the sum of the squared residuals (SSR) between them; this approach was also exploited to examine Phase 1 in (8).

Based on the variable shifts between the curves of deaths/cases detected throughout the pandemic, we adapted the definition of the WLR previously introduced (8) by alternatively considering a 1-week, a 2- week, or a 3-week shift. A regression analysis was performed on the WLR values in the weeks W_50_-W_67_ to quantify its decrease. 95% confidence intervals (95% CI) were computed according to a Poisson approximation. The significance level (0.05) of the decrease was assessed by calculating the p-value. Data were analyzed with MATLAB R2014b and R statistical software v. 4.0.0 (14,15).

The study did not involve participants and information were gathered from freely accessible public databases, and data were analyzed in aggregated form and without any identifier. Therefore, no ethical approval was required for this research. The analyses adhere to the Guidelines for Accurate and Transparent Health Estimates Reporting (GATHER) (16).

## Data Availability

All data are available

## References

1. Cascella M, Rajnik M, Aleem A, Dulebohn SC, Di Napoli R. Features, Evaluation, and Treatment of Coronavirus (COVID-19). In: StatPearls [Internet]. Treasure Island (FL): StatPearls Publishing; 2021 Apr 20.

2. Ferrara P, Albano L. COVID-19 and healthcare systems: what should we do next? Public Health 2020; 185: 1–2.

3. Muralidar S, Ambi SV, Sekaran S, Krishnan UM. The emergence of COVID-19 as a global pandemic: Understanding the epidemiology, immune response and potential therapeutic targets of SARS-CoV-2. Biochimie. 2020;179:85–100.

4. World Health Organization. WHO Coronavirus Disease (COVID-19) Dashboard. Available online: https://covid19.who.int (Last accessed on June 30, 2021).

5. COVID-19 coronavirus pandemic. Available online: https://www.worldometers.info/coronavirus/italy (Last accessed on June 30, 2021).

6. Chaqroun A, Hartard C, Schvoerer E. Anti-SARS-CoV-2 Vaccines and Monoclonal Antibodies Facing Viral Variants. Viruses 2021;13(6):1171.

7. Istituto Superiore di Sanità. Impatto della vaccinazione COVID-19 sul rischio di infezione da SARS-CoV-2 e successivo ricovero e decesso in Italia (27.12.2020 - 30.05.2021). Available online: https://www.epicentro.iss.it/vaccini/covid-19-report-valutazione-vaccinazione (Last accessed on July 4, 2021).

8. Balasco N, d’Alessandro V, Ferrara P, Smaldone G, Vitagliano L. Analysis of the time evolution of COVID-19 lethality during the first epidemic wave in Italy. Acta Biomedica 2021;92(2): e2021171.

9. Balasco N, d’Alessandro V, Smaldone G, Vitagliano L. Analysis of the time evolution of SARS-CoV-2 lethality rate in Italy: Evidence of an unaltered virus potency. doi: https://doi.org/10.1101/2020.06.12.20129387

10. Palmieri L, Palmer K, Lo Noce C, et al. Differences in the clinical characteristics of COVIDLJ19 patients who died in hospital during different phases of the pandemic: national data from Italy. Aging Clinical and Experimental Research 2021;33:193–199

11. Istituto Superiore di Sanità. Characteristics of SARS-CoV-2 patients dying in Italy (Nov 18, 2020). Available online: https://www.epicentro.iss.it/en/coronavirus/bollettino/Report-COVID-2019_18_november_2020.pdf (Last accessed on July 4, 2021).

12. Cereda D, Tirani M, Rovida F, et al. The early phase of the COVID-19 outbreak in Lombardy, Italy. 2003.09320 [q-bio.PE]

13. Dipartimento della Protezione Civile. COVID-19 Italia – Monitoraggio della situazione. Available online: www.opendatadpc.maps.arcgis.com/apps/opsdashboard/index.html#/b0c68bce2cce478eaac82fe38d4138b1 (Last accessed on June 30, 2021).

14. MATLAB R2014b, September 2014, MathWorks, Inc., Natick, Massachusetts, USA.

15. R Foundation for Statistical Computing. The R Foundation: Vienna, Austria. Available online: www.R-project.org

16. Stevens GA, Alkema L, Black RE, et al. Guidelines for Accurate and Transparent Health Estimates Reporting: the GATHER statement. Lancet 2016; 388: e19–23.

