## Supplementary figures/tables for "The temporal correlation between positive testing and death in Italy: from the first phase to the later evolution of the COVID-19 pandemic"

### Supplementary Material

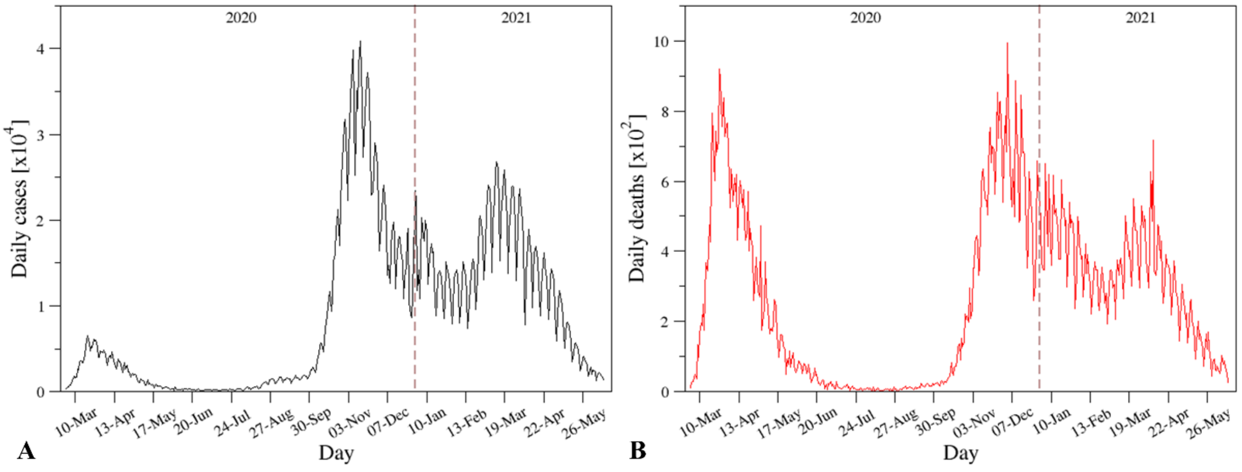


**Figure S1.** Daily evolutions of (A) cases (positive tests) and (B) deaths in the time interval March 2nd, 2020 - June 13th, 2021.

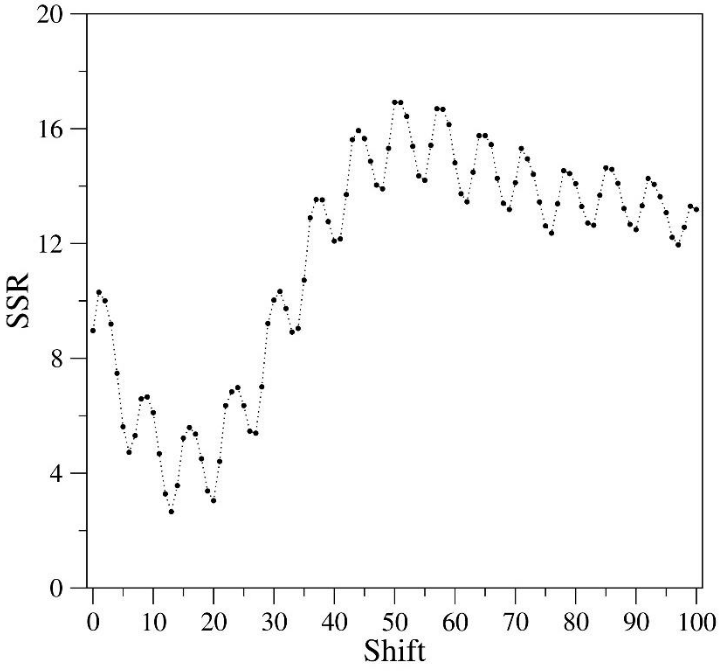


**Figure S2.** Sum of squared residuals (SSR) as a function of day shift of the curves in Phase 2 ($W_{32}$-$W_{67}$). The normalized curve of the daily cases is shifted ahead with respect to the normalized curve of the daily deaths.

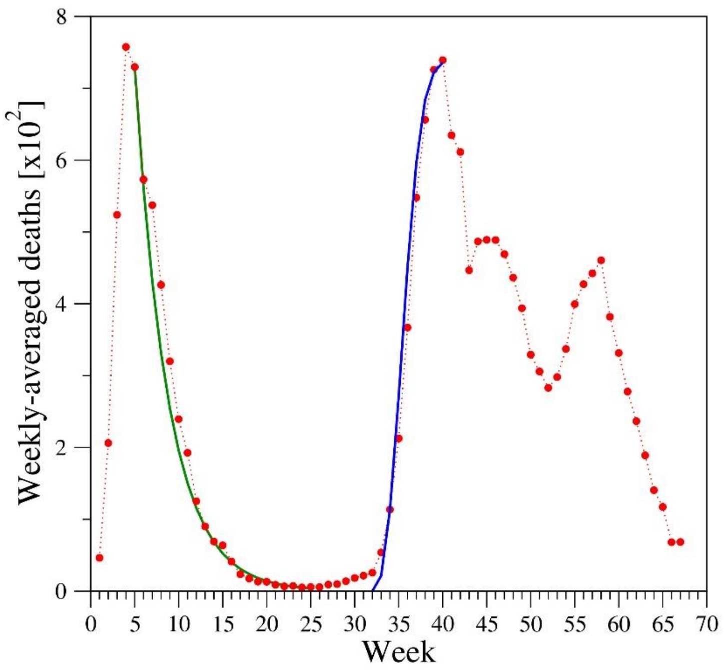


**Figure S3.** Modeling of the lockdown-driven fall of Phase 1 and the rise of Subphase 2.1 of the curve of deaths with a decreasing exponential (time constant 3.8) and a sigmoidal (exponent 2.5 and time constant 4.1) function, respectively.

**Table S1.** Daily cases, deaths, and tests in the time interval March 2nd, 2020 - June 13th, 2021 collected from the Reports of the Italian National Institute of Health (ISS).

| Date | Daily cases | Daily deaths | Daily tests |
|---|---|---|---|
| 02-Mar | 335 | 11 | 2218 |
| 03-Mar | 466 | 27 | 2511 |
| 04-Mar | 587 | 28 | 3981 |
| 05-Mar | 769 | 41 | 2525 |
| 06-Mar | 778 | 49 | 3997 |
| 07-Mar | 1247 | 36 | 5703 |
| 08-Mar | 1492 | 133 | 7875 |
| 09-Mar | 1797 | 97 | 3889 |
| 10-Mar | 1577 | 168 | 6935 |
| 11-Mar | 1713 | 196 | 12393 |
| 12-Mar | 2651 | 189 | 12857 |
| 13-Mar | 2547 | 250 | 11477 |
| 14-Mar | 3497 | 175 | 11682 |
| 15-Mar | 3590 | 368 | 15729 |
| 16-Mar | 3385 | 349 | 13063 |
| 17-Mar | 3374 | 345 | 10695 |
| 18-Mar | 4207 | 475 | 16884 |
| 19-Mar | 5322 | 427 | 17236 |
| 20-Mar | 5986 | 627 | 24109 |
| 21-Mar | 6557 | 793 | 26336 |
| 22-Mar | 5560 | 651 | 25180 |
| 23-Mar | 4790 | 601 | 17066 |
| 24-Mar | 5249 | 743 | 21496 |
| 25-Mar | 5210 | 683 | 27481 |
| 26-Mar | 6153 | 712 | 36615 |
| 27-Mar | 5909 | 919 | 33019 |
| 28-Mar | 5974 | 889 | 35447 |
| 29-Mar | 5217 | 756 | 24504 |
| 30-Mar | 4050 | 812 | 23329 |
| 31-Mar | 4053 | 837 | 29609 |
| 01-Apr | 4782 | 727 | 34455 |
| 02-Apr | 4668 | 760 | 39809 |
| 03-Apr | 4585 | 766 | 38617 |
| 04-Apr | 4805 | 681 | 37375 |
| 05-Apr | 4316 | 525 | 34237 |
| 06-Apr | 3599 | 636 | 30271 |
| 07-Apr | 3039 | 604 | 33713 |
| 08-Apr | 3836 | 542 | 51680 |
| 09-Apr | 4204 | 610 | 46244 |
| 10-Apr | 3951 | 570 | 53495 |
| 11-Apr | 4694 | 619 | 56609 |
| 12-Apr | 4092 | 431 | 46720 |

| | | | |
|---|---|---|---|
| 13-Apr | 3153 | 566 | 36717 |
| 14-Apr | 2972 | 602 | 26779 |
| 15-Apr | 2667 | 578 | 43715 |
| 16-Apr | 3786 | 525 | 60999 |
| 17-Apr | 3493 | 575 | 65705 |
| 18-Apr | 3491 | 482 | 61725 |
| 19-Apr | 3047 | 433 | 50708 |
| 20-Apr | 2256 | 454 | 41483 |
| 21-Apr | 2729 | 570 | 52126 |
| 22-Apr | 3370 | 401 | 63101 |
| 23-Apr | 2646 | 464 | 66658 |
| 24-Apr | 3021 | 420 | 62447 |
| 25-Apr | 2357 | 415 | 65387 |
| 26-Apr | 2324 | 260 | 49916 |
| 27-Apr | 1739 | 333 | 32003 |
| 28-Apr | 2019 | 382 | 57272 |
| 29-Apr | 2086 | 323 | 63827 |
| 30-Apr | 1872 | 285 | 68456 |
| 01-May | 1965 | 269 | 74208 |
| 02-May | 1900 | 474 | 55412 |
| 03-May | 1389 | 174 | 44935 |
| 04-May | 1221 | 195 | 37631 |
| 05-May | 1075 | 236 | 55263 |
| 06-May | 1444 | 369 | 64263 |
| 07-May | 1401 | 274 | 70359 |
| 08-May | 1327 | 243 | 63775 |
| 09-May | 1083 | 194 | 69171 |
| 10-May | 802 | 165 | 51678 |
| 11-May | 744 | 179 | 40740 |
| 12-May | 1402 | 172 | 67003 |
| 13-May | 888 | 195 | 61973 |
| 14-May | 992 | 262 | 71876 |
| 15-May | 789 | 242 | 68176 |
| 16-May | 875 | 153 | 69179 |
| 17-May | 675 | 145 | 60101 |
| 18-May | 451 | 99 | 36406 |
| 19-May | 813 | 162 | 63158 |
| 20-May | 665 | 161 | 67195 |
| 21-May | 642 | 156 | 71679 |
| 22-May | 652 | 130 | 75380 |
| 23-May | 669 | 119 | 72410 |
| 24-May | 531 | 50 | 55824 |
| 25-May | 300 | 92 | 35241 |
| 26-May | 397 | 78 | 57674 |
| 27-May | 584 | 117 | 67324 |

| | | | |
|---|---|---|---|
| 28-May | 593 | 70 | 75893 |
| 29-May | 516 | 87 | 72135 |
| 30-May | 416 | 111 | 69342 |
| 31-May | 333 | 75 | 54118 |
| 01-Jun | 200 | 60 | 31394 |
| 02-Jun | 319 | 55 | 52159 |
| 03-Jun | 322 | 71 | 37299 |
| 04-Jun | 177 | 88 | 49953 |
| 05-Jun | 519 | 85 | 65028 |
| 06-Jun | 270 | 72 | 72485 |
| 07-Jun | 197 | 53 | 49478 |
| 08-Jun | 280 | 65 | 27112 |
| 09-Jun | 283 | 79 | 55003 |
| 10-Jun | 202 | 71 | 62699 |
| 11-Jun | 380 | 53 | 62472 |
| 12-Jun | 163 | 56 | 70620 |
| 13-Jun | 347 | 78 | 49750 |
| 14-Jun | 337 | 44 | 56527 |
| 15-Jun | 301 | 26 | 28107 |
| 16-Jun | 210 | 34 | 46882 |
| 17-Jun | 329 | 43 | 77701 |
| 18-Jun | 332 | 66 | 58154 |
| 19-Jun | 251 | 47 | 57541 |
| 20-Jun | 264 | 49 | 54722 |
| 21-Jun | 224 | 24 | 40545 |
| 22-Jun | 221 | 23 | 28972 |
| 23-Jun | 113 | 18 | 40485 |
| 24-Jun | 190 | 30 | 53266 |
| 25-Jun | 296 | 34 | 56061 |
| 26-Jun | 255 | 30 | 52768 |
| 27-Jun | 175 | 8 | 61351 |
| 28-Jun | 174 | 22 | 37346 |
| 29-Jun | 126 | 6 | 27218 |
| 30-Jun | 142 | 23 | 48273 |
| 01-Jul | 182 | 21 | 55366 |
| 02-Jul | 201 | 30 | 53243 |
| 03-Jul | 223 | 15 | 50096 |
| 04-Jul | 235 | 21 | 52011 |
| 05-Jul | 192 | 7 | 37462 |
| 06-Jul | 208 | 8 | 22166 |
| 07-Jul | 137 | 30 | 43219 |
| 08-Jul | 193 | 15 | 50443 |
| 09-Jul | 214 | 12 | 52552 |
| 10-Jul | 276 | 12 | 47953 |

| | | | |
|---|---|---|---|
| 11-Jul | 188 | 7 | 45931 |
| 12-Jul | 234 | 9 | 38259 |
| 13-Jul | 169 | 13 | 23933 |
| 14-Jul | 114 | 17 | 41867 |
| 15-Jul | 162 | 13 | 48449 |
| 16-Jul | 230 | 20 | 50432 |
| 17-Jul | 231 | 11 | 50767 |
| 18-Jul | 249 | 14 | 48265 |
| 19-Jul | 218 | 3 | 35525 |
| 20-Jul | 190 | 13 | 24253 |
| 21-Jul | 128 | 15 | 43110 |
| 22-Jul | 280 | 9 | 49318 |
| 23-Jul | 306 | 10 | 60311 |
| 24-Jul | 252 | 5 | 53334 |
| 25-Jul | 273 | 5 | 51671 |
| 26-Jul | 252 | 5 | 40526 |
| 27-Jul | 170 | 5 | 25551 |
| 28-Jul | 181 | 11 | 48170 |
| 29-Jul | 289 | 6 | 56018 |
| 30-Jul | 382 | 3 | 61858 |
| 31-Jul | 379 | 9 | 60944 |
| 01-Aug | 295 | 5 | 60383 |
| 02-Aug | 238 | 8 | 43269 |
| 03-Aug | 159 | 12 | 24036 |
| 04-Aug | 190 | 5 | 43788 |
| 05-Aug | 384 | 10 | 56451 |
| 06-Aug | 401 | 6 | 58673 |
| 07-Aug | 552 | 3 | 59196 |
| 08-Aug | 347 | 13 | 53298 |
| 09-Aug | 463 | 2 | 37637 |
| 10-Aug | 259 | 4 | 26432 |
| 11-Aug | 412 | 6 | 40642 |
| 12-Aug | 476 | 10 | 52658 |
| 13-Aug | 522 | 6 | 51188 |
| 14-Aug | 574 | 3 | 46723 |
| 15-Aug | 627 | 4 | 53123 |
| 16-Aug | 479 | 4 | 36807 |
| 17-Aug | 320 | 4 | 30666 |
| 18-Aug | 401 | 5 | 53976 |
| 19-Aug | 642 | 7 | 71095 |
| 20-Aug | 840 | 6 | 77442 |
| 21-Aug | 947 | 9 | 71996 |
| 22-Aug | 1071 | 3 | 77674 |
| 23-Aug | 1209 | 7 | 67371 |
| 24-Aug | 952 | 4 | 45914 |

| | | | |
|---|---|---|---|
| 25-Aug | 876 | 4 | 72341 |
| 26-Aug | 1365 | 13 | 93529 |
| 27-Aug | 1409 | 5 | 94024 |
| 28-Aug | 1462 | 9 | 97065 |
| 29-Aug | 1444 | 1 | 99108 |
| 30-Aug | 1365 | 4 | 81723 |
| 31-Aug | 999 | 6 | 58518 |
| 01-Sep | 984 | 8 | 81050 |
| 02-Sep | 1332 | 6 | 102959 |
| 03-Sep | 1402 | 10 | 92790 |
| 04-Sep | 1738 | 11 | 113085 |
| 05-Sep | 1700 | 16 | 107658 |
| 06-Sep | 1303 | 7 | 76856 |
| 07-Sep | 1107 | 12 | 52553 |
| 08-Sep | 1366 | 10 | 92403 |
| 09-Sep | 1434 | 14 | 95990 |
| 10-Sep | 1597 | 10 | 94186 |
| 11-Sep | 1616 | 10 | 98880 |
| 12-Sep | 1499 | 6 | 92706 |
| 13-Sep | 1458 | 7 | 72143 |
| 14-Sep | 1008 | 14 | 45309 |
| 15-Sep | 1229 | 9 | 80517 |
| 16-Sep | 1450 | 12 | 100607 |
| 17-Sep | 1585 | 13 | 101773 |
| 18-Sep | 1906 | 10 | 99839 |
| 19-Sep | 1638 | 24 | 103223 |
| 20-Sep | 1587 | 15 | 83428 |
| 21-Sep | 1349 | 17 | 55862 |
| 22-Sep | 1392 | 14 | 87303 |
| 23-Sep | 1640 | 20 | 103696 |
| 24-Sep | 1786 | 23 | 108019 |
| 25-Sep | 1912 | 20 | 107269 |
| 26-Sep | 1869 | 17 | 104387 |
| 27-Sep | 1766 | 17 | 87714 |
| 28-Sep | 1493 | 16 | 51109 |
| 29-Sep | 1647 | 24 | 90185 |
| 30-Sep | 1851 | 19 | 105236 |
| 01-Oct | 2548 | 24 | 118236 |
| 02-Oct | 2498 | 23 | 120301 |
| 03-Oct | 2844 | 27 | 118932 |
| 04-Oct | 2578 | 18 | 92714 |
| 05-Oct | 2257 | 16 | 60241 |
| 06-Oct | 2676 | 28 | 99742 |
| 07-Oct | 3678 | 31 | 125314 |
| 08-Oct | 4458 | 22 | 128098 |

| | | | |
|---|---|---|---|
| 09-Oct | 5372 | 28 | 129471 |
| 10-Oct | 5724 | 29 | 133084 |
| 11-Oct | 5456 | 26 | 104658 |
| 12-Oct | 4616 | 39 | 85442 |
| 13-Oct | 5901 | 41 | 112544 |
| 14-Oct | 7331 | 43 | 152196 |
| 15-Oct | 8803 | 83 | 162932 |
| 16-Oct | 10010 | 55 | 150377 |
| 17-Oct | 10925 | 47 | 165837 |
| 18-Oct | 11704 | 69 | 146541 |
| 19-Oct | 9335 | 73 | 98862 |
| 20-Oct | 10874 | 89 | 144737 |
| 21-Oct | 15198 | 127 | 177848 |
| 22-Oct | 16079 | 136 | 170392 |
| 23-Oct | 19139 | 91 | 182032 |
| 24-Oct | 19644 | 151 | 177669 |
| 25-Oct | 21268 | 128 | 161880 |
| 26-Oct | 17007 | 141 | 124686 |
| 27-Oct | 21991 | 221 | 174398 |
| 28-Oct | 24989 | 205 | 198952 |
| 29-Oct | 26826 | 217 | 201452 |
| 30-Oct | 31082 | 199 | 215085 |
| 31-Oct | 31756 | 297 | 215886 |
| 01-Nov | 29907 | 208 | 183457 |
| 02-Nov | 22250 | 233 | 135731 |
| 03-Nov | 28242 | 353 | 182287 |
| 04-Nov | 30547 | 352 | 211831 |
| 05-Nov | 34498 | 428 | 219884 |
| 06-Nov | 37807 | 446 | 234245 |
| 07-Nov | 39809 | 425 | 231673 |
| 08-Nov | 32614 | 331 | 191144 |
| 09-Nov | 25263 | 356 | 147725 |
| 10-Nov | 35098 | 580 | 217758 |
| 11-Nov | 32960 | 623 | 225640 |
| 12-Nov | 37978 | 636 | 234672 |
| 13-Nov | 40896 | 550 | 254908 |
| 14-Nov | 37253 | 544 | 227695 |
| 15-Nov | 33977 | 546 | 195275 |
| 16-Nov | 27354 | 504 | 152663 |
| 17-Nov | 32188 | 731 | 208458 |
| 18-Nov | 34283 | 753 | 234834 |
| 19-Nov | 36173 | 653 | 250186 |
| 20-Nov | 37239 | 699 | 238077 |
| 21-Nov | 34767 | 692 | 237225 |
| 22-Nov | 28334 | 562 | 188747 |

| | | | |
|---|---|---|---|
| 23-Nov | 22925 | 630 | 148945 |
| 24-Nov | 23231 | 853 | 188659 |
| 25-Nov | 25851 | 722 | 230007 |
| 26-Nov | 28993 | 822 | 232711 |
| 27-Nov | 28344 | 827 | 222803 |
| 28-Nov | 26321 | 686 | 225940 |
| 29-Nov | 20647 | 541 | 130524 |
| 30-Nov | 16374 | 672 | 130524 |
| 01-Dec | 19350 | 785 | 182100 |
| 02-Dec | 20703 | 684 | 207143 |
| 03-Dec | 23236 | 993 | 220047 |
| 04-Dec | 24099 | 814 | 212741 |
| 05-Dec | 21052 | 662 | 194984 |
| 06-Dec | 18846 | 564 | 163550 |
| 07-Dec | 13612 | 528 | 111217 |
| 08-Dec | 14733 | 634 | 149232 |
| 09-Dec | 12652 | 499 | 118475 |
| 10-Dec | 16887 | 887 | 171586 |
| 11-Dec | 18550 | 761 | 190416 |
| 12-Dec | 19738 | 649 | 196439 |
| 13-Dec | 17818 | 484 | 152697 |
| 14-Dec | 11965 | 491 | 103584 |
| 15-Dec | 14714 | 846 | 164431 |
| 16-Dec | 17431 | 680 | 199489 |
| 17-Dec | 18136 | 683 | 185320 |
| 18-Dec | 17989 | 674 | 179800 |
| 19-Dec | 16306 | 553 | 176185 |
| 20-Dec | 15104 | 352 | 137420 |
| 21-Dec | 10860 | 415 | 87889 |
| 22-Dec | 13294 | 628 | 157705 |
| 23-Dec | 14521 | 553 | 183864 |
| 24-Dec | 18040 | 505 | 193777 |
| 25-Dec | 19037 | 459 | 152334 |
| 26-Dec | 10429 | 261 | 81564 |
| 27-Dec | 8909 | 305 | 59879 |
| 28-Dec | 8583 | 445 | 68681 |
| 29-Dec | 11212 | 659 | 128740 |
| 30-Dec | 16202 | 575 | 169045 |
| 31-Dec | 23476 | 555 | 186004 |
| 01-Jan | 22205 | 462 | 157524 |
| 02-Jan | 11831 | 364 | 67174 |
| 03-Jan | 14243 | 347 | 102974 |
| 04-Jan | 10797 | 348 | 77993 |
| 05-Jan | 15373 | 649 | 135106 |
| 06-Jan | 20331 | 548 | 121275 |

| | | | |
|---|---|---|---|
| 07-Jan | 18016 | 414 | 140267 |
| 08-Jan | 17531 | 620 | 172119 |
| 09-Jan | 19976 | 483 | 139758 |
| 10-Jan | 18625 | 361 | 139758 |
| 11-Jan | 12532 | 448 | 91656 |
| 12-Jan | 14241 | 616 | 141641 |
| 13-Jan | 15771 | 507 | 175429 |
| 14-Jan | 17244 | 522 | 160585 |
| 15-Jan | 16146 | 477 | 273506 |
| 16-Jan | 16309 | 475 | 261404 |
| 17-Jan | 12545 | 377 | 211078 |
| 18-Jan | 8824 | 377 | 158674 |
| 19-Jan | 10494 | 603 | 254070 |
| 20-Jan | 13548 | 524 | 279762 |
| 21-Jan | 14078 | 521 | 267567 |
| 22-Jan | 13633 | 472 | 264728 |
| 23-Jan | 13330 | 488 | 286331 |
| 24-Jan | 11627 | 299 | 216211 |
| 25-Jan | 8552 | 420 | 126931 |
| 26-Jan | 10580 | 541 | 256287 |
| 27-Jan | 15192 | 467 | 293770 |
| 28-Jan | 14361 | 492 | 275579 |
| 29-Jan | 13572 | 477 | 268750 |
| 30-Jan | 12712 | 421 | 298010 |
| 31-Jan | 11252 | 237 | 213364 |
| 01-Feb | 7916 | 329 | 142419 |
| 02-Feb | 9653 | 499 | 244429 |
| 03-Feb | 13186 | 476 | 279307 |
| 04-Feb | 13654 | 421 | 270142 |
| 05-Feb | 14215 | 377 | 270507 |
| 06-Feb | 13441 | 385 | 282407 |
| 07-Feb | 11640 | 270 | 206789 |
| 08-Feb | 7952 | 307 | 144270 |
| 09-Feb | 10621 | 422 | 274263 |
| 10-Feb | 12947 | 336 | 310994 |
| 11-Feb | 15131 | 391 | 292533 |
| 12-Feb | 13899 | 316 | 287619 |
| 13-Feb | 13524 | 311 | 290534 |
| 14-Feb | 11061 | 221 | 205642 |
| 15-Feb | 7333 | 258 | 179278 |
| 16-Feb | 10378 | 336 | 274019 |
| 17-Feb | 12067 | 369 | 294411 |
| 18-Feb | 13753 | 347 | 288458 |
| 19-Feb | 15462 | 348 | 297128 |
| 20-Feb | 14929 | 251 | 306078 |

| | | | |
|---|---|---|---|
| 21-Feb | 13439 | 232 | 250986 |
| 22-Feb | 9615 | 274 | 170672 |
| 23-Feb | 13292 | 356 | 303850 |
| 24-Feb | 16409 | 318 | 340247 |
| 25-Feb | 19875 | 308 | 353704 |
| 26-Feb | 20485 | 253 | 325404 |
| 27-Feb | 18901 | 280 | 323047 |
| 28-Feb | 17447 | 192 | 257024 |
| 01-Mar | 13094 | 246 | 170633 |
| 02-Mar | 17039 | 343 | 335983 |
| 03-Mar | 20864 | 347 | 358884 |
| 04-Mar | 22839 | 339 | 339635 |
| 05-Mar | 24028 | 297 | 378463 |
| 06-Mar | 23600 | 307 | 355024 |
| 07-Mar | 20745 | 207 | 271336 |
| 08-Mar | 13878 | 318 | 184684 |
| 09-Mar | 19615 | 376 | 345972 |
| 10-Mar | 22385 | 332 | 361040 |
| 11-Mar | 25639 | 373 | 372217 |
| 12-Mar | 26793 | 380 | 369636 |
| 13-Mar | 26051 | 317 | 372944 |
| 14-Mar | 21300 | 264 | 273966 |
| 15-Mar | 15247 | 354 | 179015 |
| 16-Mar | 20377 | 502 | 369375 |
| 17-Mar | 23025 | 431 | 369084 |
| 18-Mar | 24907 | 423 | 353737 |
| 19-Mar | 25816 | 386 | 364822 |
| 20-Mar | 23718 | 401 | 354480 |
| 21-Mar | 20149 | 300 | 277086 |
| 22-Mar | 13820 | 386 | 169196 |
| 23-Mar | 18744 | 551 | 335189 |
| 24-Mar | 21239 | 460 | 363767 |
| 25-Mar | 23798 | 460 | 349472 |
| 26-Mar | 23982 | 457 | 354982 |
| 27-Mar | 23839 | 380 | 357154 |
| 28-Mar | 19611 | 297 | 272630 |
| 29-Mar | 12954 | 417 | 156692 |
| 30-Mar | 16000 | 529 | 301451 |
| 31-Mar | 22439 | 467 | 351221 |
| 01-Apr | 23634 | 501 | 356085 |
| 02-Apr | 21918 | 481 | 331154 |
| 03-Apr | 21253 | 376 | 359214 |
| 04-Apr | 18025 | 326 | 250933 |
| 05-Apr | 10676 | 296 | 102795 |
| 06-Apr | 7745 | 421 | 112962 |

| | | | |
|---|---|---|---|
| 07-Apr | 13696 | 627 | 339939 |
| 08-Apr | 17207 | 487 | 362162 |
| 09-Apr | 18922 | 718 | 349003 |
| 10-Apr | 17558 | 344 | 334862 |
| 11-Apr | 15737 | 331 | 253100 |
| 12-Apr | 9781 | 358 | 190635 |
| 13-Apr | 13439 | 476 | 304990 |
| 14-Apr | 16157 | 469 | 334766 |
| 15-Apr | 16954 | 380 | 319633 |
| 16-Apr | 15937 | 429 | 327704 |
| 17-Apr | 15364 | 310 | 331734 |
| 18-Apr | 12693 | 251 | 230116 |
| 19-Apr | 8859 | 317 | 146728 |
| 20-Apr | 12066 | 392 | 294045 |
| 21-Apr | 13658 | 365 | 350034 |
| 22-Apr | 16229 | 361 | 364804 |
| 23-Apr | 14758 | 344 | 315700 |
| 24-Apr | 13816 | 324 | 320780 |
| 25-Apr | 13154 | 218 | 239482 |
| 26-Apr | 8438 | 303 | 145819 |
| 27-Apr | 10401 | 374 | 302734 |
| 28-Apr | 13379 | 345 | 336336 |
| 29-Apr | 14319 | 289 | 330075 |
| 30-Apr | 13445 | 264 | 338771 |
| 01-May | 12962 | 226 | 378202 |
| 02-May | 9146 | 144 | 156872 |
| 03-May | 5945 | 256 | 121829 |
| 04-May | 9110 | 305 | 315506 |
| 05-May | 10576 | 267 | 327169 |
| 06-May | 11802 | 258 | 324640 |
| 07-May | 10552 | 207 | 328612 |
| 08-May | 10173 | 224 | 338436 |
| 09-May | 8289 | 139 | 226006 |
| 10-May | 5077 | 198 | 130000 |
| 11-May | 6942 | 251 | 286428 |
| 12-May | 7849 | 262 | 306744 |
| 13-May | 8080 | 201 | 287026 |
| 14-May | 7560 | 182 | 298186 |
| 15-May | 6654 | 136 | 294686 |
| 16-May | 5752 | 93 | 202573 |
| 17-May | 3452 | 140 | 118924 |
| 18-May | 4446 | 201 | 262864 |
| 19-May | 5501 | 149 | 287256 |
| 20-May | 5738 | 164 | 251037 |
| 21-May | 5215 | 133 | 269744 |

| | | | |
|---|---|---|---|
| 22-May | 4715 | 125 | 286603 |
| 23-May | 3994 | 72 | 179391 |
| 24-May | 2486 | 110 | 107481 |
| 25-May | 3222 | 166 | 252646 |
| 26-May | 3933 | 121 | 260962 |
| 27-May | 4146 | 171 | 243967 |
| 28-May | 3738 | 126 | 249911 |
| 29-May | 3350 | 83 | 247330 |
| 30-May | 2947 | 44 | 164495 |
| 31-May | 1820 | 82 | 86977 |
| 01-Jun | 2482 | 93 | 221818 |
| 02-Jun | 2892 | 62 | 226272 |
| 03-Jun | 1967 | 59 | 97633 |
| 04-Jun | 2555 | 73 | 220939 |
| 05-Jun | 2436 | 57 | 238632 |
| 06-Jun | 2272 | 51 | 149958 |
| 07-Jun | 1271 | 65 | 84567 |
| 08-Jun | 1895 | 102 | 220917 |
| 09-Jun | 2198 | 77 | 218738 |
| 10-Jun | 2070 | 88 | 188120 |
| 11-Jun | 1900 | 69 | 217610 |
| 12-Jun | 1723 | 52 | 212966 |
| 13-Jun | 1390 | 26 | 134136 |

**Table S2.** Week definition with starting and ending date.

| Week | Starting Date | Ending Date |
|---|---:|---:|
| $W_1$ | 02/03/2020 | 08/03/2020 |
| $W_2$ | 09/03/2020 | 15/03/2020 |
| $W_3$ | 16/03/2020 | 22/03/2020 |
| $W_4$ | 23/03/2020 | 29/03/2020 |
| $W_5$ | 30/03/2020 | 05/04/2020 |
| $W_6$ | 06/04/2020 | 12/04/2020 |
| $W_7$ | 13/04/2020 | 19/04/2020 |
| $W_8$ | 20/04/2020 | 26/04/2020 |
| $W_9$ | 27/04/2020 | 03/05/2020 |
| $W_{10}$ | 04/05/2020 | 10/05/2020 |
| $W_{11}$ | 11/05/2020 | 17/05/2020 |
| $W_{12}$ | 18/05/2020 | 24/05/2020 |
| $W_{13}$ | 25/05/2020 | 31/05/2020 |
| $W_{14}$ | 01/06/2020 | 07/06/2020 |
| $W_{15}$ | 08/06/2020 | 14/06/2020 |
| $W_{16}$ | 15/06/2020 | 21/06/2020 |
| $W_{17}$ | 22/06/2020 | 28/06/2020 |
| $W_{18}$ | 29/06/2020 | 05/07/2020 |
| $W_{19}$ | 06/07/2020 | 12/07/2020 |
| $W_{20}$ | 13/07/2020 | 19/07/2020 |
| $W_{21}$ | 20/07/2020 | 26/07/2020 |
| $W_{22}$ | 27/07/2020 | 02/08/2020 |
| $W_{23}$ | 03/08/2020 | 09/08/2020 |
| $W_{24}$ | 10/08/2020 | 16/08/2020 |
| $W_{25}$ | 17/08/2020 | 23/08/2020 |
| $W_{26}$ | 24/08/2020 | 30/08/2020 |
| $W_{27}$ | 31/08/2020 | 06/09/2020 |
| $W_{28}$ | 07/09/2020 | 13/09/2020 |
| $W_{29}$ | 14/09/2020 | 20/09/2020 |
| $W_{30}$ | 21/09/2020 | 27/09/2020 |
| $W_{31}$ | 28/09/2020 | 04/10/2020 |
| $W_{32}$ | 05/10/2020 | 11/10/2020 |
| $W_{33}$ | 12/10/2020 | 18/10/2020 |
| $W_{34}$ | 19/10/2020 | 25/10/2020 |
| $W_{35}$ | 26/10/2020 | 01/11/2020 |
| $W_{36}$ | 02/11/2020 | 08/11/2020 |
| $W_{37}$ | 09/11/2020 | 15/11/2020 |
| $W_{38}$ | 16/11/2020 | 22/11/2020 |

| | | |
|---|---|---|
| $W_{39}$ | 23/11/2020 | 29/11/2020 |
| $W_{40}$ | 30/11/2020 | 06/12/2020 |
| $W_{41}$ | 07/12/2020 | 13/12/2020 |
| $W_{42}$ | 14/12/2020 | 20/12/2020 |
| $W_{43}$ | 21/12/2020 | 27/12/2020 |
| $W_{44}$ | 28/12/2020 | 03/01/2021 |
| $W_{45}$ | 04/01/2021 | 10/01/2021 |
| $W_{46}$ | 11/01/2021 | 17/01/2021 |
| $W_{47}$ | 18/01/2021 | 24/01/2021 |
| $W_{48}$ | 25/01/2021 | 31/01/2021 |
| $W_{49}$ | 01/02/2021 | 07/02/2021 |
| $W_{50}$ | 08/02/2021 | 14/02/2021 |
| $W_{51}$ | 15/02/2021 | 21/02/2021 |
| $W_{52}$ | 22/02/2021 | 28/02/2021 |
| $W_{53}$ | 01/03/2021 | 07/03/2021 |
| $W_{54}$ | 08/03/2021 | 14/03/2021 |
| $W_{55}$ | 15/03/2021 | 21/03/2021 |
| $W_{56}$ | 22/03/2021 | 28/03/2021 |
| $W_{57}$ | 29/03/2021 | 04/04/2021 |
| $W_{58}$ | 05/04/2021 | 11/04/2021 |
| $W_{59}$ | 12/04/2021 | 18/04/2021 |
| $W_{60}$ | 19/04/2021 | 25/04/2021 |
| $W_{61}$ | 26/04/2021 | 02/05/2021 |
| $W_{62}$ | 03/05/2021 | 09/05/2021 |
| $W_{63}$ | 10/05/2021 | 16/05/2021 |
| $W_{64}$ | 17/05/2021 | 23/05/2021 |
| $W_{65}$ | 24/05/2021 | 30/05/2021 |
| $W_{66}$ | 31/05/2021 | 06/06/2021 |
| $W_{67}$ | 07/06/2021 | 13/06/2021 |

**Table S3.** Cases and deaths *per* week of Phase 1 (blue, $W_1$-$W_{17}$) and Phase 2 (green, $W_{32}$-$W_{67}$). Average daily values were obtained dividing the total weekly number of cases/deaths by seven.

| **Week** | **Average number of cases** | **Average number of deaths** |
|---|---|---|
| $W_1$ | 811 | 46 |
| $W_2$ | 2482 | 206 |
| $W_3$ | 4913 | 524 |
| $W_4$ | 5500 | 758 |
| $W_5$ | 4466 | 730 |
| $W_6$ | 3916 | 573 |
| $W_7$ | 3230 | 537 |
| $W_8$ | 2672 | 426 |
| $W_9$ | 1853 | 320 |
| $W_{10}$ | 1193 | 239 |
| $W_{11}$ | 909 | 193 |
| $W_{12}$ | 632 | 125 |
| $W_{13}$ | 448 | 90 |
| $W_{14}$ | 286 | 69 |
| $W_{15}$ | 285 | 64 |
| $W_{16}$ | 273 | 41 |
| $W_{17}$ | 203 | 24 |
| $W_{18}$ | 186 | 18 |
| $W_{19}$ | 207 | 13 |
| $W_{20}$ | 196 | 13 |
| $W_{21}$ | 240 | 9 |
| $W_{22}$ | 276 | 7 |
| $W_{23}$ | 357 | 7 |
| $W_{24}$ | 478 | 5 |
| $W_{25}$ | 776 | 6 |
| $W_{26}$ | 1268 | 6 |
| $W_{27}$ | 1351 | 9 |
| $W_{28}$ | 1440 | 10 |
| $W_{29}$ | 1486 | 14 |
| $W_{30}$ | 1673 | 18 |
| $W_{31}$ | 2208 | 22 |
| $W_{32}$ | 4232 | 26 |
| $W_{33}$ | 8470 | 54 |
| $W_{34}$ | 15934 | 114 |
| $W_{35}$ | 26223 | 213 |
| $W_{36}$ | 32252 | 367 |
| $W_{37}$ | 34775 | 548 |
| $W_{38}$ | 32905 | 656 |
| $W_{39}$ | 25187 | 726 |
| $W_{40}$ | 20523 | 739 |

| | | |
|---|---|---|
| $W_{41}$ | 16284 | 635 |
| $W_{42}$ | 15949 | 611 |
| $W_{43}$ | 13584 | 447 |
| $W_{44}$ | 15393 | 487 |
| $W_{45}$ | 17236 | 489 |
| $W_{46}$ | 14970 | 489 |
| $W_{47}$ | 12219 | 469 |
| $W_{48}$ | 12317 | 436 |
| $W_{49}$ | 11958 | 394 |
| $W_{50}$ | 12162 | 329 |
| $W_{51}$ | 12480 | 306 |
| $W_{52}$ | 16575 | 283 |
| $W_{53}$ | 20316 | 298 |
| $W_{54}$ | 22237 | 337 |
| $W_{55}$ | 21891 | 400 |
| $W_{56}$ | 20719 | 427 |
| $W_{57}$ | 19460 | 442 |
| $W_{58}$ | 14506 | 461 |
| $W_{59}$ | 14332 | 382 |
| $W_{60}$ | 13220 | 332 |
| $W_{61}$ | 11727 | 278 |
| $W_{62}$ | 9492 | 237 |
| $W_{63}$ | 6845 | 189 |
| $W_{64}$ | 4723 | 141 |
| $W_{65}$ | 3403 | 117 |
| $W_{66}$ | 2346 | 68 |
| $W_{67}$ | 1778 | 68 |

**Table S4.** Weekly lethality rate (WLR) values with 95% Confidence Intervals (95%CI). The WLR values were calculated applying a 1-week, 2-week, and 3-week shift between positive test and death in the time intervals $W_1$-$W_{33}$, $W_{34}$-$W_{41}$, and $W_{42}$-$W_{67}$, respectively. WLR values were calculated by dividing the average daily number of deaths of a given week ($W_i$) by the average daily number of cases of one week ($W_{i-1}$), two weeks ($W_{i-2}$), or three weeks ($W_{i-3}$) before.

| **Week** | **WLR 1-week shift** | | **Week** | **WLR 2-week shift** | | **Week** | **WLR 3-week shift** | |
|---|---|---|---|---|---|---|---|---|
| | | *95% CI* | | | *95% CI* | | | *95% CI* |
| $W_2$ | 25.43 | (24.1 – 26.8) | $W_{34}$ | 2.68 | (2.5 – 2.9) | $W_{42}$ | 2.43 | (2.4 – 2.5) |
| $W_3$ | 21.10 | (20.4 – 21.8) | $W_{35}$ | 2.51 | (2.4 – 2.6) | $W_{43}$ | 2.18 | (2.1 – 2.3) |
| $W_4$ | 15.42 | (15.0 – 15.8) | $W_{36}$ | 2.30 | (2.2 – 2.4) | $W_{44}$ | 2.99 | (2.9 – 3.1) |
| $W_5$ | 13.27 | (12.9 – 13.6) | $W_{37}$ | 2.09 | (2.0 – 2.2) | $W_{45}$ | 3.07 | (3.0 – 3.2) |
| $W_6$ | 12.83 | (12.4 – 13.2) | $W_{38}$ | 2.03 | (2.0 – 2.1) | $W_{46}$ | 3.60 | (3.5 – 3.7) |
| $W_7$ | 13.72 | (13.3 – 14.1) | $W_{39}$ | 2.09 | (2.0 – 2.1) | $W_{47}$ | 3.05 | (2.9 – 3.2) |
| $W_8$ | 13.20 | (12.7 – 13.7) | $W_{40}$ | 2.25 | (2.2 – 2.3) | $W_{48}$ | 2.53 | (2.4 – 2.6) |
| $W_9$ | 11.98 | (11.5 – 12.5) | $W_{41}$ | 2.52 | (2.4 – 2.6) | $W_{49}$ | 2.63 | (2.5 – 2.7) |
| $W_{10}$ | 12.92 | (12.3 – 13.6) | | | | $W_{50}$ | 2.69 | (2.6 – 2.8) |
| $W_{11}$ | 16.14 | (15.3 – 17.0) | | | | $W_{51}$ | 2.48 | (2.4 – 2.6) |
| $W_{12}$ | 13.78 | (12.9 – 14.7) | | | | $W_{52}$ | 2.37 | (2.3 – 2.5) |
| $W_{13}$ | 14.24 | (13.2 – 15.4) | | | | $W_{53}$ | 2.45 | (2.3 – 2.6) |
| $W_{14}$ | 15.42 | (14.1 – 16.9) | | | | $W_{54}$ | 2.70 | (2.6 – 2.8) |
| $W_{15}$ | 22.25 | (20.2 – 24.4) | | | | $W_{55}$ | 2.41 | (2.3 – 2.5) |
| $W_{16}$ | 14.51 | (12.9 – 16.3) | | | | $W_{56}$ | 2.10 | (2.0 – 2.2) |
| $W_{17}$ | 8.63 | (7.4 – 10.1) | | | | $W_{57}$ | 1.99 | (1.9 – 2.1) |
| $W_{18}$ | 8.64 | (7.2 – 10.3) | | | | $W_{58}$ | 2.10 | (2.0 – 2.2) |
| $W_{19}$ | 7.15 | (5.8 – 8.8) | | | | $W_{59}$ | 1.84 | (1.8 – 1.9) |
| $W_{20}$ | 6.28 | (5.1 – 7.7) | | | | $W_{60}$ | 1.70 | (1.6 – 1.8) |
| $W_{21}$ | 4.52 | (3.5 – 5.8) | | | | $W_{61}$ | 1.92 | (1.8 – 2.0) |
| $W_{22}$ | 2.80 | (2.1 – 3.7) | | | | $W_{62}$ | 1.65 | (1.6 – 1.7) |
| $W_{23}$ | 2.64 | (2.0 – 3.5) | | | | $W_{63}$ | 1.43 | (1.4 – 1.5) |
| $W_{24}$ | 1.48 | (1.0 – 2.0) | | | | $W_{64}$ | 1.20 | (1.1 – 1.3) |
| $W_{25}$ | 1.22 | (0.9 – 1.7) | | | | $W_{65}$ | 1.24 | (1.2 – 1.3) |
| $W_{26}$ | 0.74 | (0.5 – 1.0) | | | | $W_{66}$ | 1.00 | (0.9 – 1.1) |
| $W_{27}$ | 0.72 | (0.6 – 0.9) | | | | $W_{67}$ | 1.45 | (1.3 – 1.6) |
| $W_{28}$ | 0.73 | (0.6 – 0.9) | | | | | | |
| $W_{29}$ | 0.96 | (0.8 – 1.2) | | | | | | |
| $W_{30}$ | 1.23 | (1.0 – 1.5) | | | | | | |
| $W_{31}$ | 1.29 | (1.1 – 1.5) | | | | | | |
| $W_{32}$ | 1.16 | (1.0 – 1.3) | | | | | | |
| $W_{33}$ | 1.27 | (1.1 – 1.4) | | | | | | |
